# The Impact of Distractions on Intracortical Brain-Computer Interface Control of a Robotic Arm

**DOI:** 10.1101/2021.01.28.21250556

**Authors:** Michael D. Guthrie, Angelica J. Herrera, John E. Downey, Lucas J. Brane, Michael L. Boninger, Jennifer L. Collinger

## Abstract

This was an investigational device observational trial with the objective to evaluate the impact of distractions on intracortical brain-computer interface (BCI) performance. Two individuals with tetraplegia had microelectrode arrays implanted into their motor cortex for trials of intracortical BCI safety and performance. The primary task was moving a robotic arm between two targets as quickly as possible, performed alone and with various secondary distraction conditions. Primary outcomes included targets acquired, path efficiency, and subjective difficulty. There was no difference in the number of targets acquired for either subject with or without distractions. Median path efficiency was similar across all conditions (range: 0.766-0.846) except the motor distraction for Subject P2, where the median path efficiency dropped to 0.675 (p = 0.033, Mann-Whitney U test). Both subjects rated the overall difficulty of the task with and without distractions as low. Overall, intracortical BCI performance was robust to various distractions.

## Introduction

Many medical conditions, including spinal cord injuries, can lead to a devastating loss of upper limb function resulting in significant disability. Restoring hand and arm function is a priority for people with tetraplegia due to cervical spinal cord injury^1, 2^. Rehabilitation aims to restore movement capabilities and functional abilities; however, motor deficits often persist^3^. When recovery has plateaued, assistive technologies can be used to augment or replace function. Brain-computer interfaces (BCIs) are one type of assistive device that use intact brain activity to control an end effector such as a robotic arm or computer cursor^4, 5^. Intracortical BCIs use microelectrode arrays to record activity from 10s-100s of neurons, commonly from the motor cortex where the firing rate patterns of these neurons provides detailed information about movement intention^6, 7^. People with tetraplegia have been able to use intracortical BCIs to interact with a computer^8, 9^, to control robotic devices that replace reaching and grasping function^10, 11^, and to restore grasping function of their own hand using functional electrical stimulation^12, 13^. Surveys of populations of potential BCI users, including those with spinal cord injuries, suggest that ease of use and reliable device performance are important design features^2, 14, 15^. Therefore, an ideal BCI would be simple to use and capable of restoring functional abilities in a consistent and reliable manner. To date, the majority of intracortical BCI research has occurred in a controlled, laboratory setting. However, an important step towards clinical translation is to demonstrate robust performance in real world environments where distractions may be present.

Everyday use of a BCI would require the system to output reliable, continuous control in the face of external stimuli such as environmental disturbances or fluctuations in mental states. Previous studies on attention and distraction in BCIs have focused primarily on electroencephalography (EEG)-based interfaces that record neural activity through electrodes placed on the scalp. EEG BCIs typically rely on motor imagery (e.g. imagining moving your right hand to move a cursor to the right) or sensory evoked potentials to operate the interface^4, 16^. Research on attention in EEG-based BCI performance suggests that in able-bodied individuals, there is an optimal level of attention and cognitive activity that results in more accurate BCI performance^17, 18^. Additional experiments have sought to characterize the influence of specific types of distractions, such as simple visual and auditory distractions on performance and have found that these types of distractions do not significantly impair EEG-based BCI systems^19-21^. However, various studies have demonstrated that distractions with increasing cognitive demand led to detectable impairments in performance of EEG-based BCIs^22, 23^. A proposed mechanism of these performance impairments is that attentional shifts during distraction attenuate the activity recorded from motor cortex that is used to control the BCI^24^. Ideally, BCI users will be able to successfully use a system while engaged in other activities. Evidence of impaired performance in EEG-based BCI in the presence of distractions poses a significant barrier to the practical application of BCIs as an assistive technology.

Another barrier to the successful translation of BCI use into real-world environments is that some individuals may require significant training and learning to achieve optimal performance^25^. Others may not be able to learn how to operate a BCI at all, which is termed BCI illiteracy^26^. However, in many cases intracortical BCIs have enabled high performance control with minimal training or learning^27-29^. Despite the potential for practical and functional use of intracortical BCIs, the impact of distractions on intracortical BCI performance has not been studied directly and it is unclear if intracortical BCI would be more robust to cognitive distractions than EEG-based systems. One recent study demonstrated that intracortical BCI control of a computer cursor was not impaired when the user was simultaneously speaking^30^. This is particularly significant because neural activity in the hand knob area of motor cortex recorded with the BCI was modulated during stand-alone speech; however, during BCI control, the patterns of neural activity associated with speech did not overlap with those used for BCI control. To inform the design and functional use of intracortical BCI systems it will be important to further characterize precisely which types of distraction may disrupt the typical performance of the system. For example, given that single neurons in motor cortex can be modulated by many types of movements^31^, it is possible that the performance of simultaneous movement-based tasks may interfere with BCI control. The aim of the current study is to characterize the impact of multiple types of distractions on intracortical BCI performance during robotic arm control. We hypothesize that BCI performance will be robust to simple auditory distractions, tasks designed to increase cognitive load, and speech-related activities, while simultaneous movement-based tasks will lead to reduced performance.

## Materials and Methods

### Participants

Two participants completed this study as a part of a clinical trial conducted under Investigational Device Exemptions granted by the United States Food and Drug Administration and registered at clinicaltrials.gov (NCT01364480 and NCT01894802). Informed consent was obtained from both participants prior to the completion of any study-related procedures. All procedures followed protocol and accord with the ethical standards the Space and Naval Warfare System Center Pacific. Ethical approval was granted by the University of Pittsburgh Institutional Review Board.

Subject P1 was woman in her 50’s with spinocerebellar degeneration resulting in motor complete tetraplegia at the C4 level with some preserved sensation^32^. Two 96-channel intracortical microelectrode arrays^a^ were implanted in the hand and arm region of her left motor (M1) cortex. Data were collected over 5 study sessions, occurring between 595- and 609-days post-implant. Subject P2 was a male in his 20’s with tetraplegia due to a C5 AIS B spinal cord injury^33^. Subject P2 had two 88-channel intracortical microelectrode arrays^a^ implanted in the hand and arm region of his left motor (M1) cortex. He participated in 4 study sessions, occurring between 606- and 626-days post-implant. Subject P2 also had arrays implanted into somatosensory cortex for experiments regarding restoring sensation^34^ though those were not used for the experiments presented here. Both participants completed a variety of BCI tasks, including robotic arm control, prior to these experiments and had demonstrated skilled reach and grasp control using the BCI without distractions^35^. Study visits were typically completed 3 days per week for 3-4 hours per day for the duration of the implant.

### Neural recording and BCI decoder calibration

Neural data was collected using the Neuroport Neural Signal Processor^a^. A threshold for all recorded channels was set at −5.25 times root-mean-square voltage (RMS) for Subject P1 and −4.5 times RMS for Subject P2 at the beginning of each test session. Spike counts, identified by threshold crossings, were binned for each channel every 30ms (33 Hz update rate) for Subject P1 and every 20ms (50 Hz update rate) for Subject P2. Binned spike counts were low-pass filtered using an exponential smoothing function with a 450ms and 440ms window for Subjects P1 and P2, respectively, and were square-root transformed.

Both subjects utilized a modular prosthetic limb^b^ throughout the experimental sessions. To provide the participants with BCI control of the robotic arm, a decoder was trained to transform neural firing rates into three-dimensional endpoint translation velocity commands. As presented in our previous studies, an indirect optimal linear estimation (OLE) decoder was trained in virtual reality (VR) using a two-step calibration method^7^. In the first step, the participants were instructed to attempt to move a VR robotic arm to a specified target location while the computer controlled the kinematics of the VR arm. After 36-60 trials, an OLE decoder was created to predict three-dimensional endpoint velocity (v_x_, v_y_, v_z_) of the robotic hand from the recorded neural firing rates^36, 37^. Using this decoder, the participants then performed the same VR task with assisted BCI control, where the computer attenuated command signals orthogonal to the target direction^38^. After another 36-60 trials, a new decoder was trained using the neural and kinematic data collected during assisted BCI use. This decoder was used to enable BCI control of the robotic arm for the remainder of the testing session.

### BCI performance during distractions

At the beginning of each trial, the robotic arm was placed in the center of the workspace between two foam targets that were 0.4m apart (Figure 1). The participants were instructed to use the BCI to move the robotic arm as quickly as possible to alternate between the two targets. The primary task was to acquire as many targets as possible in 60 seconds. A target was deemed to be acquired when the robotic hand made contact with the intended physical target on the appropriate end of the workspace. If contact was made with the same target twice in a row, it was considered only a single target acquisition.

**Figure 1.**
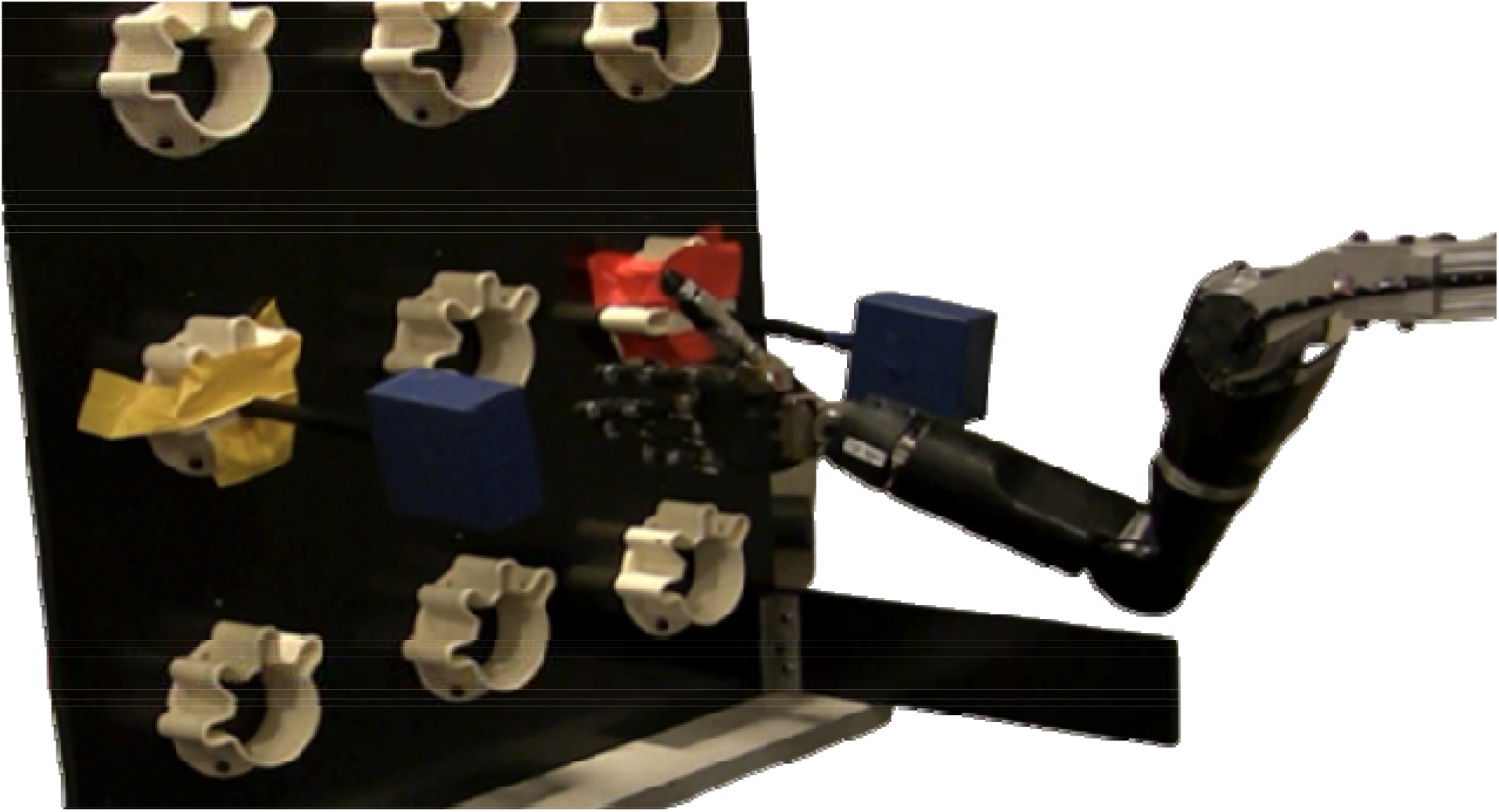
Experimental setup. The robotic arm was placed in between two foam targets (blue). The participants were seated to the left of the robotic prosthesis while using the BCI to control the endpoint velocity of the hand.

During an experimental session, two 60-second trials were collected for each condition. The six task conditions were: No Distraction (Video 1 -https://drive.google.com/file/d/19n4lombhCrq7rklhU8mrjJcco-lN7xCD/view?usp=sharing), Background Noise (Video 2 -https://drive.google.com/file/d/1RXJkalXGWL1vkXqdk1nHHyaVRbBFqt8V/view?usp=sharing), Counting Tones (Video 3 -https://drive.google.com/file/d/1zJrz55KsNupGQBTMWmBrCf5j29FTNIju/view?usp=sharing), Counting Back by Three’s (Video 4 -https://drive.google.com/file/d/1FjzH1A92dp1F9Pa1h_pOaDxbEGwBkY2H/view?usp=sharing), Casual Conversation (Video 5 -https://drive.google.com/file/d/1x4LxXBsyLMJRMeS8KkNhw2zeLXTxOoei/view?usp=sharing), and a Motor task (Video 6 -https://drive.google.com/file/d/1BCMDuhaImNr2vLe-D9Q0iMt9duVaTsyR/view?usp=sharing). The No Distraction condition was used as a basis of comparison for the distraction conditions. Further details of each distraction condition are provided in Table 1. Trial order was randomized for each session. The background noise condition was designed to mimic the auditory conditions of a noisy environment outside of the laboratory, specifically a restaurant. Prior research in psychology and BCI performance has made use of distractions with high cognitive load, or internal thought processes that require the use of working memory^39, 40^. We included a tone counting task, which is a variant of an auditory n-back test^41, 42^, as well as a counting back task^43^ to assess how cognitive distraction may impact BCI performance. Casual conversation included the cognitive effort of listening to and responding to conversational questions as well as the motor task of speaking. Finally, each subject performed an overt movement task that was compatible with their specific injury level and was distinct from the BCI task. Subject P2 was limited to a single trial for each condition during one testing day due to time constraints. Subject P2 also attempted to manipulate a wheelchair joystick with his hand as a motor distraction task, however had significant shoulder pain related to the movement. It is unclear if the movement itself or the associated pain caused large changes in neural firing rates, but this prevented him from completing the primary task and this data was not included in the analysis. Instead, we selected a movement with a smaller range of motion that he would commonly do, swiping across a tablet with his right hand. This was introduced in later sessions, so he completed fewer overall trials of the motor task.

**Table 1.** Distraction conditions. Explanation of the various conditions including auditory, cognitive, and motor distractions.

| Condition | Description |  |
| --- | --- | --- |
| No Distraction | The primary task was completed without any secondary distractions. |  |
| Background Noise | Subjects listened to an audio track with background restaurant noise at a constant volume. |  |
| Counting Tones | Beeps with low, middle, and high-frequency tones were presented randomly and at variable time intervals to the subjects. The subjects were instructed to add the number of high beeps, subtract the number of low beeps, and count the value out loud throughout the trial. |  |
| Counting Back by Three's | The researcher generated a random three-digit number and the subjects were instructed to count backward by three's out loud starting from the generated number. |  |
| Casual Conversation | The subjects were asked questions (which were never repeated) in order to mimic normal conversation. Example questions included:<br>*How was your weekend?*<br>*Did you watch any movies yesterday?* |  |
| Motor Task | Subject P1, Wheelchair operation:<br>The subject's power wheelchair was turned off during the trials. The subject received an auditory cue every 10 seconds to change the direction of force she was applying to the chin joystick. She applied force throughout the entire trial. The direction moved clockwise from a random starting direction (up, down, left, or right). | Subject P2, Tablet swiping:<br>The subject was instructed to replicate the motion of swiping a tablet with his right hand in response to a tone every 10 seconds throughout the duration of the trial. |

Performance measures included the total number of targets acquired per 60-second trial during back-and-forth movements of the robotic arm, the three-dimensional path efficiency of the robotic arm between targets, and the user-reported subjective difficulty of each trial (on a 10-point scale where 1 = very easy and 10 = very difficult). Path efficiency was calculated as the ratio of the ideal path length to actual path length so that a direct movement towards a target would have a path efficiency of 1.0. The actual path length was calculated as the three-dimensional distance traveled between each successful acquisition of the right and left targets (defined as the point of direction change between reaches) based on the robotic arm endpoint position feedback. Since successful target acquisition was achieved by touching any location on the physical target (Figure 1), we calculated the ideal path length for each trial by finding the average location each target was acquired by the robotic hand and determining the distance between these two points. The kinematic data from four trials (3 for Subject P1, including Background Noise, Counting Back by Three’s, and Casual Conversation trials and 1 for Subject P2, No Distraction trial) was not recorded properly. This occurred in rare cases where one of the robotic fingers became displaced preventing position feedback from being saved, although the commanded velocities of the arm and online control were not impacted. These trials were omitted from the kinematic analysis but included for the number of targets acquired and subjective difficulty ratings. The performance metrics were tested for normality using the Lillefors test^44^ and we determined that non-parametric tests would be appropriate for all outcome measures. Statistical analyses were completed separately for each subject. We evaluated whether performance during each of the independent distraction conditions was significantly different that the No Distraction condition using the Mann-Whitney U test. A significance level of 0.05 was used, and we did not include an adjustment for multiple comparisons, as Type 1 errors were acceptable given that we were looking for any small impairments in BCI performance.

## Results

Both participants were able to use the BCI to control the robotic arm to complete the primary target acquisition task, which involved active control of the trajectory of the arm with frequent direction changes. Subject P1 acquired a median of 20 targets (interquartile range (IQR): 16.5 – 21) per 60 second trial across all conditions and Subject P2 acquired a median of 23 targets (IQR: 18 – 26) across all conditions. There were no significant differences in the number of targets acquired during the distraction conditions as compared to the No Distraction condition for either subject (Mann-Whitney U tests, Figure 2).

**Figure 2.**
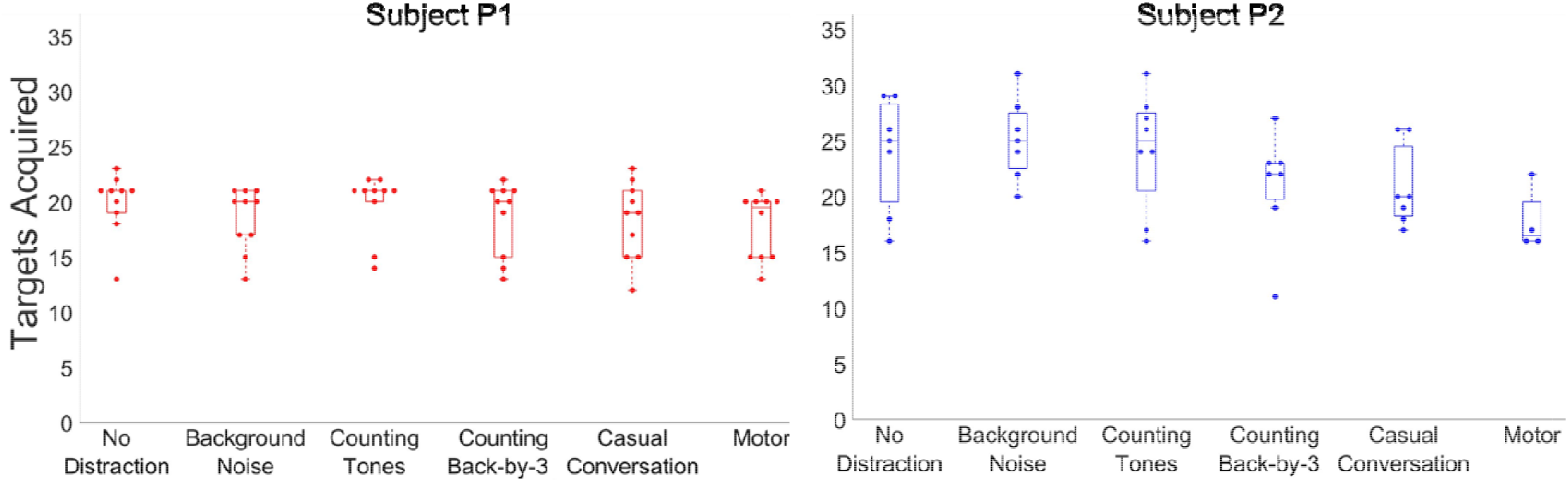
Targets Acquired for each Distraction Condition. The number of targets acquired for each 60-second trial is displayed for Subject P1 (left, red dots) and Subject P2 (right, blue dots) for each condition. Boxes represent the 25^th^ percentile, median, and 75^th^ percentile. Dashed lines extend to values that are outside the interquartile range and are not outliers. Neither subject demonstrated a significant difference in the number of targets acquired during any of the distractions as compared to the No Distraction condition.

In addition to acquiring the targets in a timely manner, a BCI should enable precise control of the robotic arm position during the movements, which would be reflected as a high path efficiency (i.e., closer to 1.0). Both subjects had similar median path efficiencies and ranges across all trials, 0.814 (IQR: 0.772 – 0.847) for Subject P1 and 0.819 (IQR: 0.762– 0.856) for Subject P2 (Figure 3). The only significant drop in performance was in the motor distraction condition for Subject P2 (p = 0.033, Mann Whitney U test).

**Figure 3.**
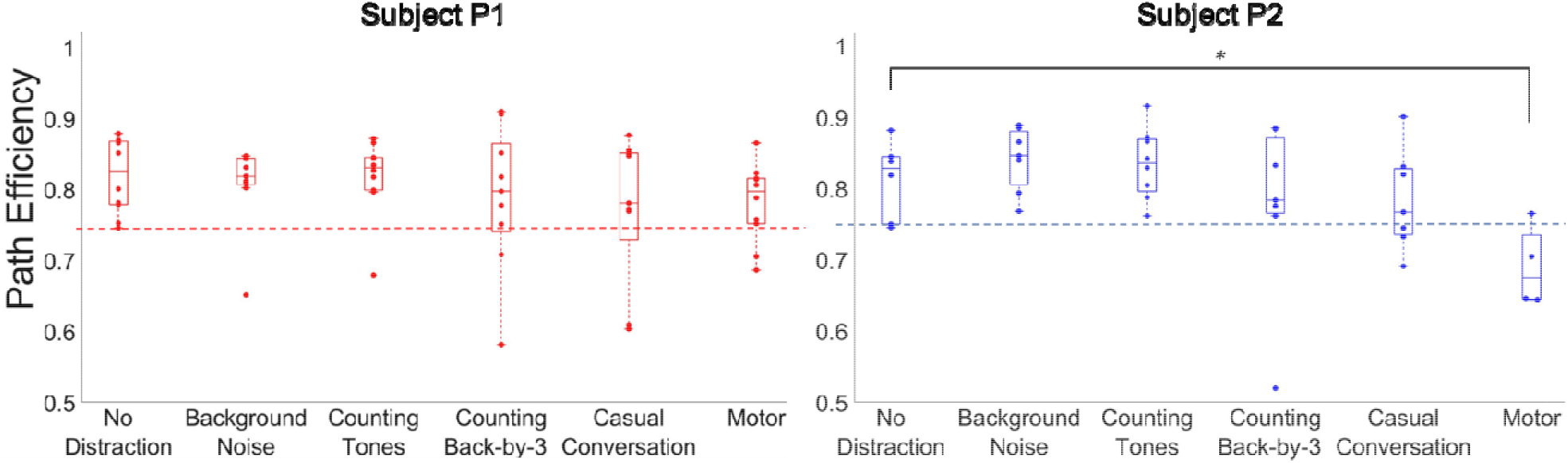
Path Efficiency for each Distraction Condition. Path efficiencies, calculated as the average of all lateral movements in a single trial, are displayed for each trial condition for Subject P1 (left, red dots) and Subject P2 (right, blue dots). Boxes outline the 25^th^ percentile, median, and 75^th^ percentile. Dashed vertical lines extend to values that are outside the interquartile range and are not outliers. The dashed horizontal line indicates the lowest path efficiency for the No Distraction condition for each Subject. ^*^Indicates statistically significant difference between the distraction condition and No Distraction for Subject P2 (Mann-Whitney U test)

There were 8 distraction trials for Subject P1 and 7 distraction trials for Subject P2 where the path efficiency dropped below that of the lowest No Distraction trial. This was chosen as a threshold for an impaired trial because it reflected the lower limit of performance for the BCI system without distraction. The lowest path efficiency for both Subject P1 and Subject P2 in the No Distraction condition was approximately 0.745. Notably, 7 out of 8 impaired trials occurred on the same testing day for Subject P1. Three out of the 4 impaired trials for Subject P2 were motor distraction trials, and these were spread across multiple testing sessions. Representative kinematic tracings of the path of the robotic arm for Subject P1 are displayed in Figure 4 for three different trial cases: a typical No Distraction trial, a typical Counting Back by Three’s trial, and an impaired Counting Back by Three’s trial.

**Figure 4.**
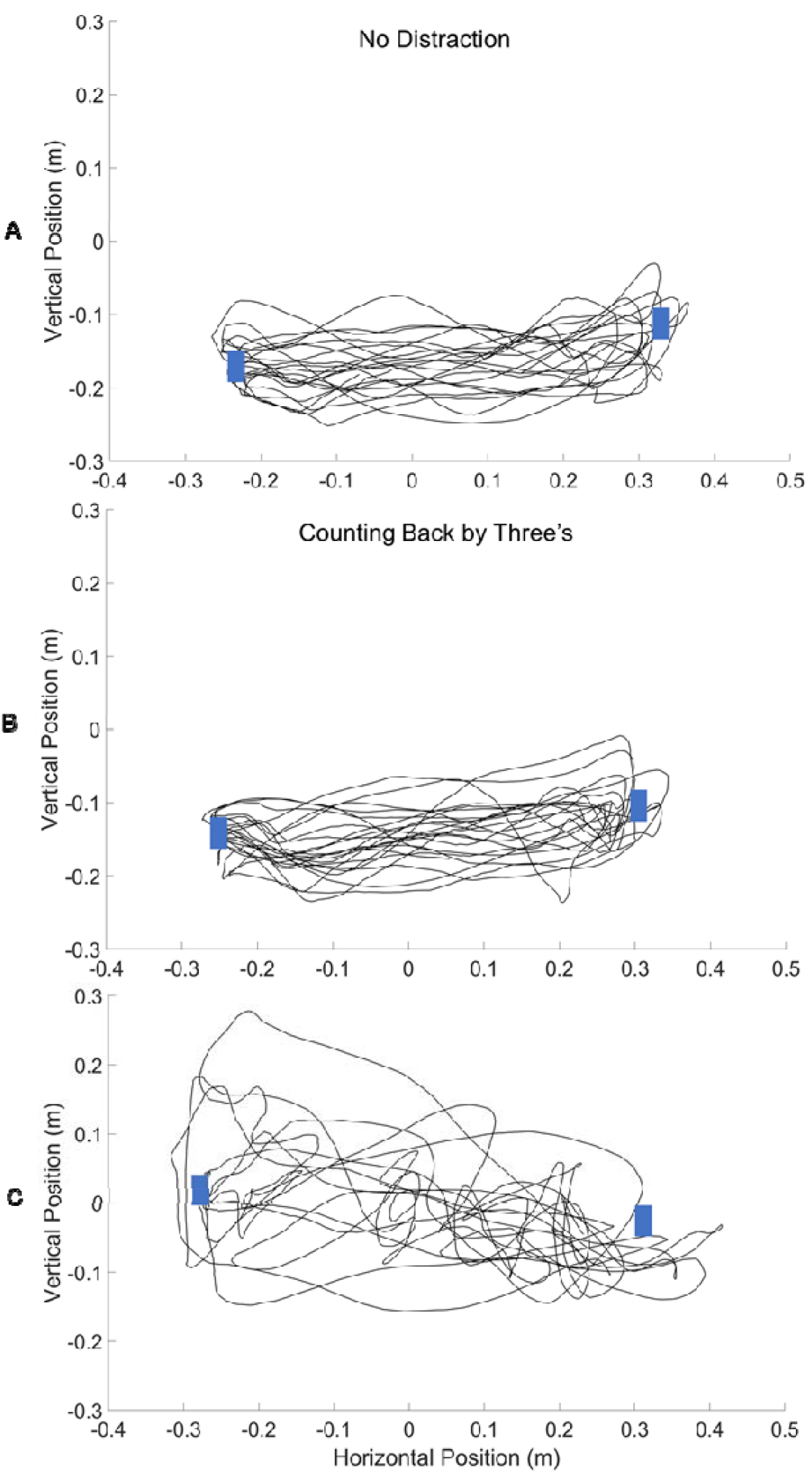
Kinematic Trajectories. The kinematic tracings of the path of the robotic arm in the vertical and horizontal plane are demonstrated for Subject P1 completing a trial with No Distraction (A, path efficiency = 0.868), Counting Back by Three’s with good path efficiency (B, path efficiency = 0.851), and Counting Back by Three’s with poor path efficiency (C, path efficiency = 0.581). The blue rectangles indicate the average position of the acquired target on either side.

Subjective difficulty ratings were collected to characterize the user perspective on how challenging it was to operate the BCI system across the various conditions (Figure 5). Subject P1 reported a median subjective difficulty of 3 (IQR: 3 - 4) for the No Distraction condition and 3 (IQR: 3 - 5) across all distraction tasks. Subject P2 rated the No Distraction condition as 1 in difficulty for all trials, compared to a median rating of 2 across all distraction conditions (IQR: 1- 3). Subject P2 did report a significantly higher median difficulty rating for the Counting Tones (p = 0.009), Counting Back by Three’s (p < 0.001), Casual Conversation (p = 0.008), and Motor distraction (p = 0.002) tasks when compared to the No Distraction condition (Mann-Whitney U test). Overall, both subjects considered the task difficulty to be very low.

**Figure 5.**
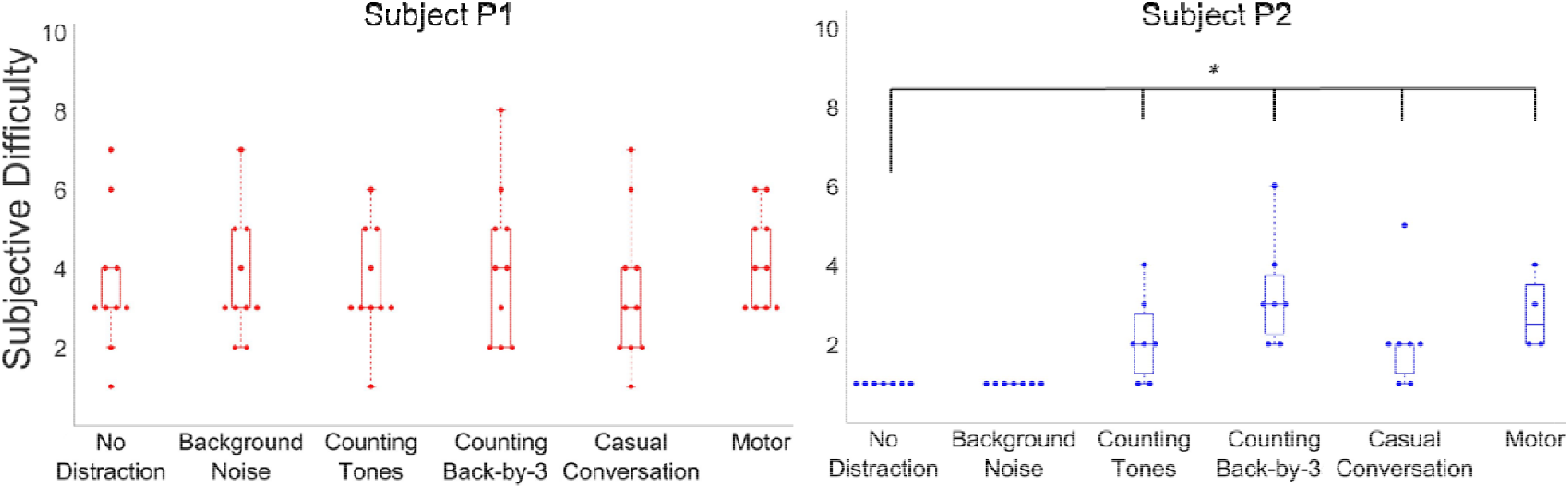
Subjective Difficulty Ratings for each Distraction Condition. The difficulty ratings on a 10-point scale (1 = very easy, 10 = very difficult) for Subject P1 (left, red dots) and Subject P2 (right, blue dots) are displayed for each trial. Boxes represent the 25^th^ percentile, median, and 75^th^ percentile. Dashed lines extend to values that are outside the interquartile range and are not outliers. ^*^ Indicates statistically significant difference between the distraction condition and No Distraction for Subject P2 (p < 0.05, Mann-Whitney U test)

## Discussion

### Overall performance

The purpose of the study was to determine the influence of various types of distraction on intracortical BCI performance. Both subjects demonstrated the ability to complete a primary BCI-controlled task in the presence of distractions. Consistent with our hypothesis, distractions overall did not significantly impair intracortical BCI performance in two subjects as determined by the number of targets acquired. Similarly, median path efficiency was unaffected with one exception; it was reduced during the Motor task in Subject P2. Median difficulty ratings were low for both subjects for all distraction conditions, although for Subject P2 this was at times significantly higher than for BCI control without distraction.

### Distractions with Increasing Cognitive Load

As expected, listening to background noise did not impair performance or subjectively increase the level of difficulty for either subject. This is consistent with findings from a previous EEG-based BCI study where there was no change in performance observed when subjects simply listened to a recording of counting numbers^21^. While listening to background noise alone is a passive activity, a BCI user may desire to maintain control of the system while actively attending to other cognitive tasks. As portable intracortical BCI systems are emerging as a potential assistive technology^45^, it will be important for individuals to be able to use their BCI in environments outside of a controlled laboratory setting. To evaluate the impact of distractions that would require cognitive effort we used two tasks, Tone Counting and Counting Back by Three’s, that have previously been shown to impair performance in a virtual hand grasping task^41^ and standing balance in older adults^43^. There was no impairment in performance measures for either subject in these two tasks, and the increase in subjective difficulty reported by Subject P2 for the cognitive tasks was modest and likely does not reflect a clinically significant barrier to the use of the system. This is opposed to previous work in EEG-based BCI, which demonstrated impaired performance in the presence of distractions with high cognitive load^22^. In some of the EEG-based studies, there were identifiable changes in the neural activity recorded during distraction conditions which impacted the resulting performance of the BCI^23, 24^. The obvious difference between the different types of BCIs is that intracortical interfaces decode neural signals from very small populations of neurons in contrast to EEG-based interfaces that record signals from the scalp surface, leading to lower spatial resolution and generally noisier signals. A simple explanation for the robustness of intracortical BCIs is that the motor neurons being recorded by the implanted array are less affected by cognitive distraction conditions, which require attention from distinct cortical areas such as the prefrontal cortex^46^. The current findings suggest that intracortical BCIs will function predictably during the intended task even in the presence of distractions requiring cognitive effort.

### Casual Conversation

Speech involves both cognitive processes as well as the execution of specific motor movements that may impair motor cortical activity used for BCI control. Indeed, a previous EEG-BCI study demonstrated that simultaneous speech during control of a virtual prosthetic arm impaired performance^22^. Single unit studies have demonstrated broad activity patterns related to movements of all four limbs and the face in the hand knob area of motor cortex^31^. In addition, prior intracortical neural research demonstrated that specific primary motor neurons were active when participants simply read a verb or imagined doing an action^47^. Despite concerns that speech may invoke cognitive or motor activity that would disrupt consistent control of a BCI, our results indicate that intracortical BCI control is not impaired by speech-related activities as both subjects were able to perform the primary task while holding a casual conversation with good performance and ease of use. A previous intracortical BCI study demonstrated that speech modulated neural activity in areas thought to be specific to the hand/arm area of motor cortex. However, despite the speech-related neural activity, the authors found that these signals did not impair BCI performance during cursor control while the participant was speaking. Neural data demonstrated that speech-related activity in the motor cortex region of interest was attenuated during BCI control enabling robust decoding of movement-related activity^30^. This explanation is also consistent with our findings.

### Motor-related Distractions

Finally, we wanted to test whether intracortical BCI performance would be maintained while performing a simultaneous movement-based task. As Subject P1 had 0/5 muscle strength in all four extremities^32^, but regularly operated her wheelchair via chin movements to control a joystick, we chose this movement to be her motor distraction. Subject P2 had trace 1/5 strength movements of right wrist extension which he would commonly use for swiping a tablet, thus this was used as a motor distraction for him. Similar to the speech condition, it would be expected that the neural activity generated to enable chin movement would be independent from that used for BCI control of a robotic arm and hand. Indeed, Subject P1 was not significantly impaired during her motor distraction. In contrast, hand movements generated by Subject P2 during tablet swiping did indeed impair performance as most path efficiencies during the motor task trial were lower than the efficiencies observed in the No Distraction trial. Observation of his motor trials does indeed reveal that fluctuations in the trajectory of the robotic prosthetic arm occurred immediately after he swiped his right hand (Video 7 - https://drive.google.com/file/d/1-KQhcEPlmux_A-_fYErKzb3Z7lFsABF1/view?usp=sharing). This implies that the neural activity used for BCI control was disrupted when he executed movement of his contralateral hand. This is consistent with results from non-human primate work that showed that BCI control was more impaired when the primate was required to decouple neural activity used for BCI control from that used to generate wrist movement of the contralateral limb^48^. These findings and results from our motor distraction trials suggest that while some motor movements may not influence intracortical BCI performance, certain specific movements may indeed disrupt the system in a predictable way. This extends our understanding of the limitations of motor cortical activity used for intracortical BCIs and informs the design of functional devices.

### Reliability and Clinical Relevance

It is intuitively expected that distractions will impair task performance. For example, use of a cell phone while driving is clearly linked to an increased risk of automobile accidents^49^. A survey of individuals with spinal cord injuries found that a majority of respondents preferred BCIs that would consistently perform with at least 80% accuracy^15^. To identify small changes in performance of the BCI system related to distraction, we used a relatively strict threshold for inaccuracy defined as any path efficiency lower than observed during the No Distraction condition. Subject P1 completed 42/50 distraction trials (84%) and Subject P2 completed 26/33 distraction trials (78.8%) with a path efficiency at least as high as the lowest No Distraction trial. It should be noted the trials with lower path efficiency were not impaired in terms of the total number of targets acquired. Also, despite occasionally higher difficulty ratings, neither subject consistently rated the task as difficult to complete with any of the distractions. Nonetheless, the inefficient trials should be addressed as they may put the technology at risk for abandonment^50^ and raise concern regarding the use of a BCI-controlled prosthetic for tasks that require significant reliability, such as picking up a glass of water. Interestingly, all but one of the impaired trials for Subject P1 occurred during the same session (the first day of testing), which generally had a lower number of targets acquired and higher difficulty ratings than the other days. The lack of any obvious technical or internal factors unique to this session may suggest a rapid learning effect for this subject. Our study helps to identify which specific types of distractions are the most challenging, and perhaps which distractions would be amenable to early detection and augmentation of the system for safe operation, similar to existing automatic safety mechanisms in the automobile industry to avoid collisions^51, 52^. Future directions of intracortical BCI research should aim to characterize the neural activity that occurs during distraction to develop methods that are robust to these changes, thus improving overall safety and reliability. While our results support that the intracortical BCI system was easy to use and performed well even in the presence of distractions, we also identified a need to detect occasional impairments in performance that may occur.

### Study Limitations

A limitation of our study is that we only conducted the experiments with two subjects due to the inherent nature of the research and interventions involving an implanted BCI. These participants were experienced with using a BCI, so our findings do not generalize to novice BCI users who may be more susceptible to distractions. Additionally, the primary task of repetitively moving a prosthetic arm back and forth between targets is relatively simple. BCI tasks that require more cognitive effort or working memory may also be more susceptible to distractions^39, 40^. Furthermore, we did not use an objective measure of cognitive load or distraction, but rather subjective difficulty ratings to gauge the user’s perspective. User reported limitations and functionality should be assessed more completely in future trials. This may include testing primary functional tasks with the BCI system that the user personally hopes to perform as well as specific conditions he or she may face in their daily use of a BCI.

### Conclusions

Two subjects with intracortical electrodes were able to successfully control a robotic prosthetic arm to complete a motor task with consistent performance in the presence of various distractions. This supports the practical use of an intracortical BCI in patients with tetraplegia to restore upper extremity function in a realistic setting. These findings are in contrast to previous EEG-based BCI distraction studies in which BCI classification accuracy and movement trajectory were impaired by distractions. Despite generally good performance, there were occasional trials where the BCI system performed below the baseline level expected for each subject. This highlights a need to detect occasional changes in neural activity and BCI performance to ensure safe and reliable BCI performance.

## Data Availability

Specific datasets can be made available upon request

## Acknowledgements

We would like to acknowledge the study participants and their families for their cooperation and effort throughout the duration of the study. We would also like to extend a special thanks to Gina McKernan for her assistance with statistical analyses and Lynne Yash for photo editing.

## Abbreviations

(BCI): Brain-computer interface
(EEG): electroencephalography
(RMS): root-mean-square voltage
(OLE): optimal linear estimation
(VR): virtual reality
(IQR): interquartile range

## Suppliers

a. NeuroPort intracortical microelectrode arrays and Neural Signal Processor: Blackrock Microsystems LLC 630 Komas Drive, Suite 200 Salt Lake City, UT 84108-1229
b. Modular Prosthetic Limb: Johns Hopkins Applied Physics Laboratory 11100 Johns Hopkins Road Laurel, Maryland 20723

